# The Robust Bidirectional Association Between Chronic Lung Disease and Incident Osteoporosis: A Two-Stage Individual Participant Data Meta-Analysis of Three International Longitudinal Cohorts (HRS, SHARE, and ELSA)

**DOI:** 10.64898/2026.03.18.26348689

**Authors:** Jinying Bao, Daming Jiang

## Abstract

**Background:** The association between chronic lung disease (CLD) and osteoporosis (OP) is well-recognized, but the direction and magnitude of this relationship remain debated, particularly in aging populations. We aimed to quantify the bidirectional association between CLD (including COPD and asthma) and incident OP using a two-stage individual participant data (IPD) meta-analysis of three large longitudinal cohorts.

**Methods:** We harmonized and analyzed individual-level data from the Health and Retirement Study (HRS, USA), the Survey of Health, Ageing and Retirement in Europe (SHARE, Europe), and the English Longitudinal Study of Ageing (ELSA, UK), all comprising adults aged ≥50 years. In the first stage, Cox proportional hazards models were fitted separately in each cohort to estimate hazard ratios (HRs) for the forward (CLD→OP) and reverse (OP→CLD) associations, adjusting for a comprehensive set of confounders (demographics, lifestyle, comorbidities, functional status). In the second stage, cohort-specific log HRs were pooled using fixed-effect meta-analysis. Heterogeneity was assessed with the I^2^ statistic.

**Results:** A total of 40,050 participants were included across the three cohorts. The pooled HR for incident OP among individuals with baseline CLD was 1.37 (95% confidence interval [CI] 1.24–1.51), with similar estimates for COPD (HR 1.47, 95% CI 1.27–1.69) and asthma (HR 1.35, 95% CI 1.22–1.50). For the reverse association, baseline OP was associated with increased risk of incident CLD (pooled HR 1.16, 95% CI 1.05–1.29), COPD (HR 1.28, 95% CI 1.11–1.47), and asthma (HR 1.17, 95% CI 1.05–1.30). Heterogeneity was low across all analyses (I^2^ ≤7.5%).

**Conclusion:** This two-stage IPD meta-analysis provides robust evidence of a bidirectional relationship between CLD and OP in older adults. These findings underscore the need for integrated screening and management of both conditions in aging populations.

**Mini Abstract:** This two-stage individual participant data meta-analysis of 40,050 participants from three international longitudinal studies demonstrates a robust bidirectional association between chronic lung disease and incident osteoporosis, supporting integrated clinical management.

## Introduction

The pathological link between CLD and OP is complex and multi-factorial. CLD, particularly COPD, is associated with systemic inflammation, characterized by elevated circulating levels of pro-inflammatory cytokines such as interleukin-6 (IL-6) and tumor necrosis factor-alpha (TNF-a), which are known to stimulate osteoclast activity and inhibit osteoblast function, leading to bone loss[1]. Furthermore, common risk factors such as smoking, physical inactivity, and vitamin D deficiency contribute to both conditions [2]. While the “forward” association (CLD -> OP) is largely attributed to these shared risk factors and disease-specific mechanisms (e.g., corticosteroid use in asthma/COPD), the “reverse” association (OP -> CLD) is less understood but mechanistically plausible. Osteoporosis-related vertebral compression fractures (VCFs) can lead to thoracic kyphosis, reduced chest wall compliance, and restricted lung volume, potentially contributing to the development or exacerbation of CLD symptoms[3].

Despite the established biological plausibility, the true nature of the bidirectional association remains a subject of controversy. Most existing studies are cross-sectional, which, while indicating a strong association, cannot distinguish between cause and effect, often leading to an overestimation of the effect size due to reverse causation bias [4]. Longitudinal studies, which are essential for inferring temporality, have yielded inconsistent results, particularly regarding the reverse association (OP -> CLD) [5]. This inconsistency is further complicated by differences in cohort characteristics, diagnostic criteria, and the extent of covariate adjustment across studies conducted in various geographical regions [4, 5].

To overcome the limitations of single-cohort studies and the heterogeneity in existing literature, we conducted a robust multi-cohort Meta-analysis. This study leverages three major international longitudinal cohorts—the Health and Retirement Study (HRS, US), the Survey of Health, Ageing and Retirement in Europe (SHARE, Europe), and the English Longitudinal Study of Ageing (ELSA, UK)—to provide a high-level, pooled estimate of the bidirectional association between CLD and incident OP[6]. Furthermore, we included the Longitudinal Ageing Study in India (LASI) as a cross-sectional comparison to empirically quantify the degree of effect size inflation inherent in cross-sectional designs. By strictly adhering to the STROBE guidelines for observational studies, our findings aim to provide the most reliable evidence to date, clarifying the complex bidirectional relationship and informing clinical guidelines for the integrated management of CLD and OP in the global aging population.

## Methods

### Study Design and Data Sources

This study employed a two-stage individual participant data (IPD) meta-analysis of three prospective population-based longitudinal cohorts: the Health and Retirement Study (HRS, USA), the Survey of Health, Ageing and Retirement in Europe (SHARE, Europe), and the English Longitudinal Study of Ageing (ELSA, UK). All cohorts are harmonized members of the HRS international family of aging studies, enabling consistent variable definitions and analytical approaches. Additionally, we included the Longitudinal Ageing Study in India (LASI) as a cross-sectional comparator to empirically evaluate the inflation of effect estimates inherent in cross-sectional designs.

Ethical approval for each original study was obtained by the respective institutional review boards, and all participants provided written informed consent. The present study involved secondary analysis of fully anonymized publicly available data; therefore, no additional ethical approval was required.

### Study Population

All cohorts comprised community-dwelling adults aged 50 years or older at baseline. For each cohort, we applied identical inclusion and exclusion criteria:

**Inclusion criteria:** age ≥50 years at baseline; complete data on exposure, outcome, and all covariates.

**Exclusion criteria:** For the forward association analysis (CLD → incident OP), participants with prevalent OP at baseline were excluded. For the reverse association analysis (OP → incident CLD), participants with prevalent CLD at baseline were excluded.

The final analytical samples were as follows:

**HRS:** 15,643 participants (forward) and 14,463 participants (reverse); median follow-up 7.5 years. **SHARE:** 18,425 participants (forward) and 19,282 participants (reverse); median follow-up 6.0 years. **ELSA:** 5,521 participants (forward) and 4,878 participants (reverse); median follow-up 13.7 years.

**LASI:** 52,951 participants (cross-sectional).

Participant selection flowcharts for each cohort are presented in Figure 1 (A-D).

**Figure 1.**
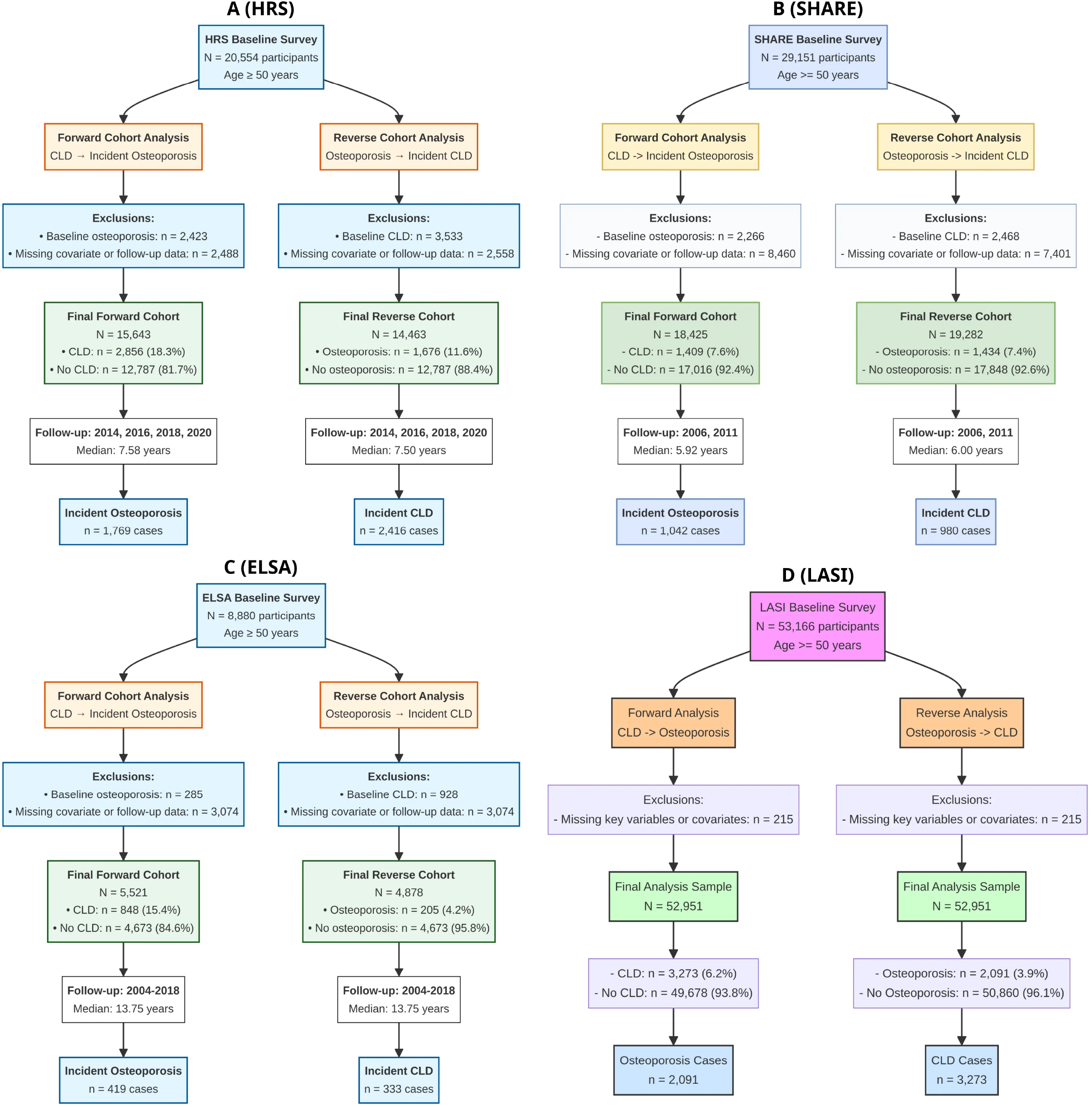
Flowchart of Participant Selection for the Four Cohorts. (A) HRS Cohort Selection. (B) SHARE Cohort Selection. (C) ELSA Cohort Selection. (D) LASI Cohort Selection.

### Variables and Harmonization

All variables were harmonized across cohorts to ensure consistency in definitions and measurement.

#### Exposure variables

Chronic lung disease (CLD) was defined as a self-reported physician diagnosis of either chronic obstructive pulmonary disease (COPD) or asthma at baseline. COPD and asthma were also examined separately as secondary exposures. All exposures were treated as binary variables.

#### Outcome variables

In the longitudinal cohorts (HRS, SHARE, ELSA), incident OP or incident CLD (COPD/asthma) was defined as the first self-reported physician diagnosis of the condition during follow-up among participants free of the outcome at baseline. Time to event was measured in years from baseline interview to the date of first diagnosis or censoring (loss to follow-up, death, or end of study). In the cross-sectional LASI cohort, outcomes were prevalent OP or CLD at the time of survey.

#### Covariates

A comprehensive set of potential confounders was selected based on prior literature and clinical plausibility. These included:

- Demographics: age (continuous), sex (male/female), marital status (married/partnered vs. other), education (categorical, harmonized across cohorts)
- Lifestyle: smoking status (current/former/never), alcohol drinking (current/former/never), physical activity level (categorical)
- Anthropometrics: body mass index (BMI, continuous, kg/m^2^)
- Comorbidities: hypertension, diabetes, heart disease, stroke, cancer, arthritis (all binary)
- Functional status: activities of daily living (ADL) score (continuous), depression risk (binary, based on validated scales)

#### Missing data

Participants with missing data for any exposure, outcome, or covariate were excluded from the primary analysis (complete case analysis). The proportion of excluded participants and reasons for exclusion are detailed in the flowcharts (Figure 1).

### Statistical Analysis

#### First stage: cohort-specific analyses

For each longitudinal cohort (HRS, SHARE, ELSA), we fitted separate Cox proportional hazards models to estimate the bidirectional associations between CLD and incident OP. Specifically:

- Forward association: CLD (and separately COPD, asthma) as exposure, incident OP as outcome
- Reverse association: OP as exposure, incident CLD (and separately COPD, asthma) as outcome

All models were adjusted for the full set of covariates listed above. The proportional hazards assumption was tested using Schoenfeld residuals and was satisfied for all models. Results were reported as hazard ratios (HRs) with 95% confidence intervals (CIs).

For the cross-sectional LASI cohort, we used multivariable logistic regression to estimate odds ratios (ORs) with 95% CIs for the corresponding associations, adjusting for the same covariates.

#### Second stage: IPD meta-analysis

Cohort-specific log HRs and their standard errors from the three longitudinal cohorts were pooled using a fixed-effect meta-analysis with the inverse variance method. We chose a fixed-effect model a priori because:

- All cohorts were derived from the same harmonized family of aging studies
- The research question was directional (testing a specific hypothesis)
- Heterogeneity was expected to be low based on prior knowledge

Heterogeneity was assessed using the I^2^ statistic, with I^2^ <25% considered low, 25–50% moderate, and >50% high. Pooled estimates were presented as HRs with 95% CIs. Forest plots were generated to visualize individual cohort estimates and pooled results.

All statistical analyses were performed using R version 4.2.1 (R Foundation for Statistical Computing, Vienna, Austria) with the survival, meta, and mice packages. A two-sided P-value <0.05 was considered statistically significant.

## Data Availability

The data underlying this article are available from the Health and Retirement Study (HRS), the Survey
of Health, Ageing and Retirement in Europe (SHARE), the English Longitudinal Study of Ageing
(ELSA), and the Longitudinal Ageing Study in India (LASI). All data are publicly available upon
request from the respective study websites.

https://releases.sharedataportal.eu/

https://hrsdata.isr.umich.edu/user/password

https://beta.ukdataservice.ac.uk/myaccount/login?referrer=%2fmyaccount

https://lasi-india.org/

## Acknowledgments

The authors would like to thank all the participants of the Health and Retirement Study (HRS), the Survey of Health, Ageing and Retirement in Europe (SHARE), the English Longitudinal Study of Ageing (ELSA), and the Longitudinal Ageing Study in India (LASI) for their invaluable contributions. We also acknowledge the dedicated efforts of the research teams and funding agencies responsible for the design, data collection, and maintenance of these important studies.

## Ethical Considerations

All original studies (HRS, SHARE, ELSA, and LASI) were conducted in accordance with the Declaration of Helsinki and were approved by their respective institutional review boards or ethics committees. All participants provided written informed consent prior to enrollment. The present study involved secondary analysis of fully anonymized, publicly available data; therefore, no additional ethical approval or informed consent was required. This manuscript does not contain any individual person’s data in any form.

## Data availability

The data underlying this article are available from the Health and Retirement Study (HRS), the Survey of Health, Ageing and Retirement in Europe (SHARE), the English Longitudinal Study of Ageing (ELSA), and the Longitudinal Ageing Study in India (LASI). All data are publicly available upon request from the respective study websites.

## Results

### Baseline Characteristics and Flowchart

The final analytical samples and exclusion processes for each cohort are detailed in Figure 1 (A-D). Baseline characteristics of the four cohorts are presented in Table 1. Across the three longitudinal cohorts (HRS, SHARE, and ELSA), the prevalence of CLD at baseline ranged from 15.4% (ELSA) to 18.3% (HRS), while OP prevalence ranged from 4.9% (ELSA) to 13.5% (HRS). The cross-sectional LASI cohort exhibited distinctly different characteristics, with substantially lower BMI (mean 22.49 kg/m^2^ vs. 26.41–28.61 kg/m^2^ in Western cohorts) and lower OP prevalence (3.9% vs. 4.9–13.5%). Detailed baseline characteristics stratified by CLD subgroups for each cohort are provided in Supplementary Tables 1–4.

**Table 1.**
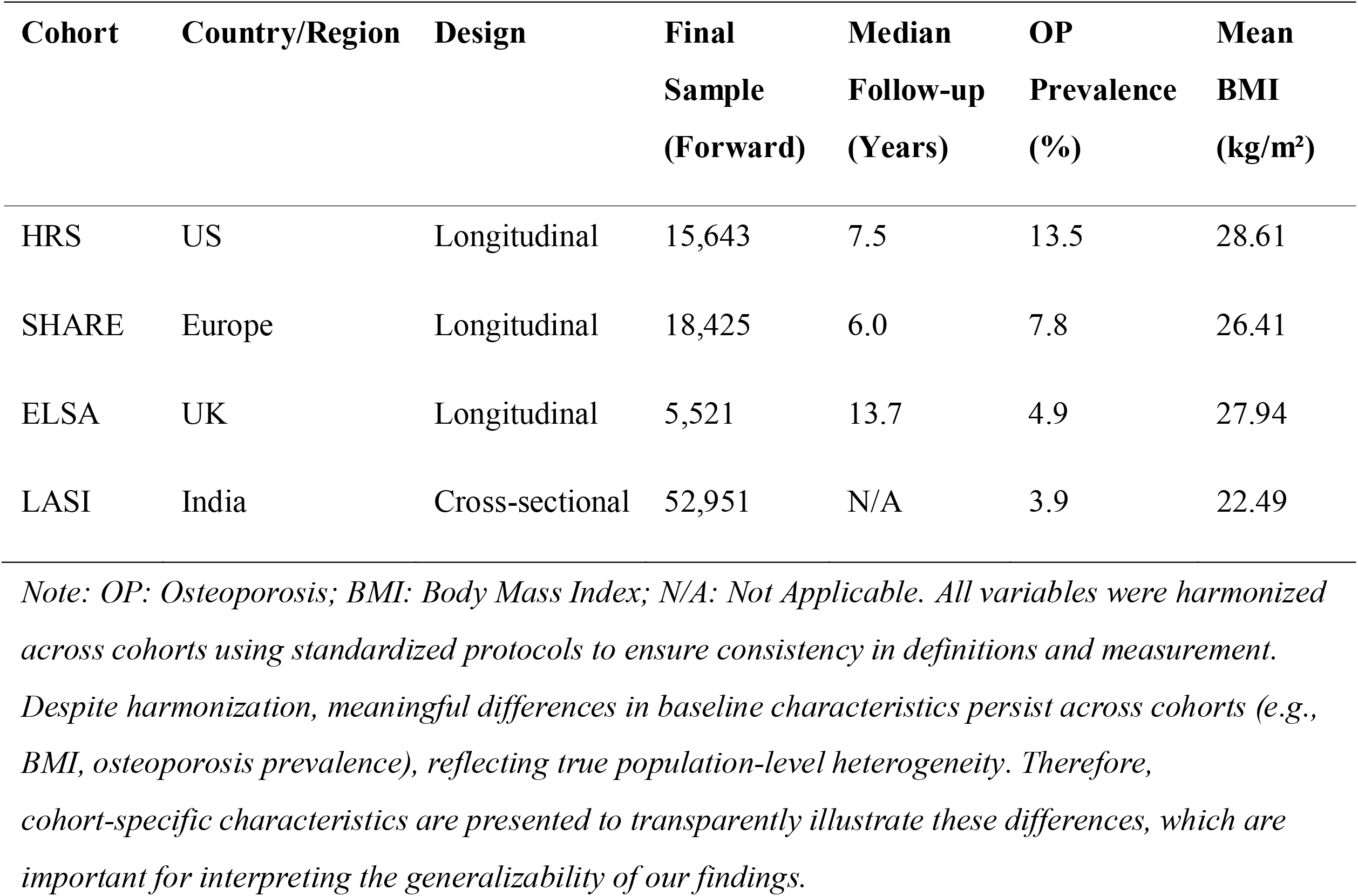
Summary of Baseline Characteristics and Cohort Design.

### Longitudinal Cohort Analysis

Table 2 summarizes the cohort-specific associations between CLD and incident OP. In HRS, baseline CLD was associated with a 35% increased risk of incident OP (HR 1.35, 95% CI 1.19–1.53), with similar estimates for COPD (HR 1.45, 95% CI 1.20–1.75) and asthma (HR 1.34, 95% CI 1.19–1.51). For the reverse association, baseline OP was associated with a 28% increased risk of incident COPD (HR 1.28, 95% CI 1.07–1.54) and a 16% increased risk of incident asthma (HR 1.16, 95% CI 1.03–1.30), though the association with overall CLD did not reach statistical significance (HR 1.12, 95% CI 0.99–1.26, P=0.058).

**Table 2.**
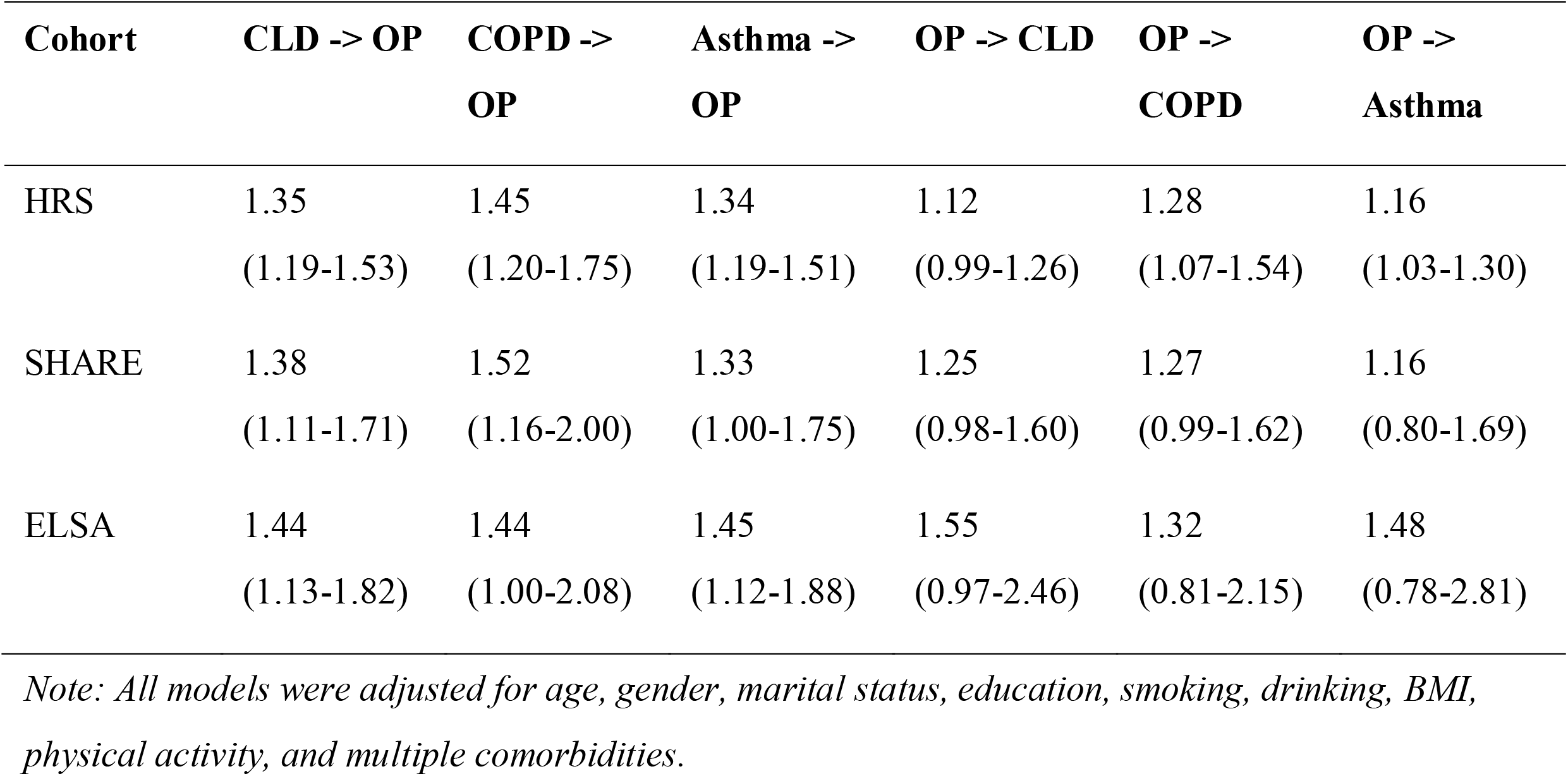
Bidirectional Association in Individual Longitudinal Cohorts (HR, 95% CI)

In SHARE, CLD was associated with a 38% increased risk of incident OP (HR 1.38, 95% CI 1.11–1.71), with stronger associations for COPD (HR 1.52, 95% CI 1.16–2.00) than asthma (HR 1.33, 95% CI 1.00–1.75). The reverse associations were directionally consistent but did not achieve statistical significance (all P>0.05), likely reflecting limited statistical power.

In ELSA, despite the smallest sample size, consistent patterns emerged. Baseline CLD was associated with a 44% increased risk of incident OP (HR 1.44, 95% CI 1.13–1.82), with similar estimates for COPD (HR 1.44, 95% CI 1.00–2.08) and asthma (HR 1.45, 95% CI 1.12–1.88). The reverse associations showed elevated but non-significant HRs (e.g., OP→CLD: HR 1.55, 95% CI 0.97–2.46), consistent with the smaller number of OP cases at baseline (n=205).

Detailed results for all analyses, including full covariate adjustments, are provided in Supplementary Tables 5–7.

### Meta-Analysis of Longitudinal Cohorts

The two-stage IPD meta-analysis pooled estimates from the three longitudinal cohorts, providing robust evidence for bidirectional associations (Table 3, Figure 2).

**Table 3.**
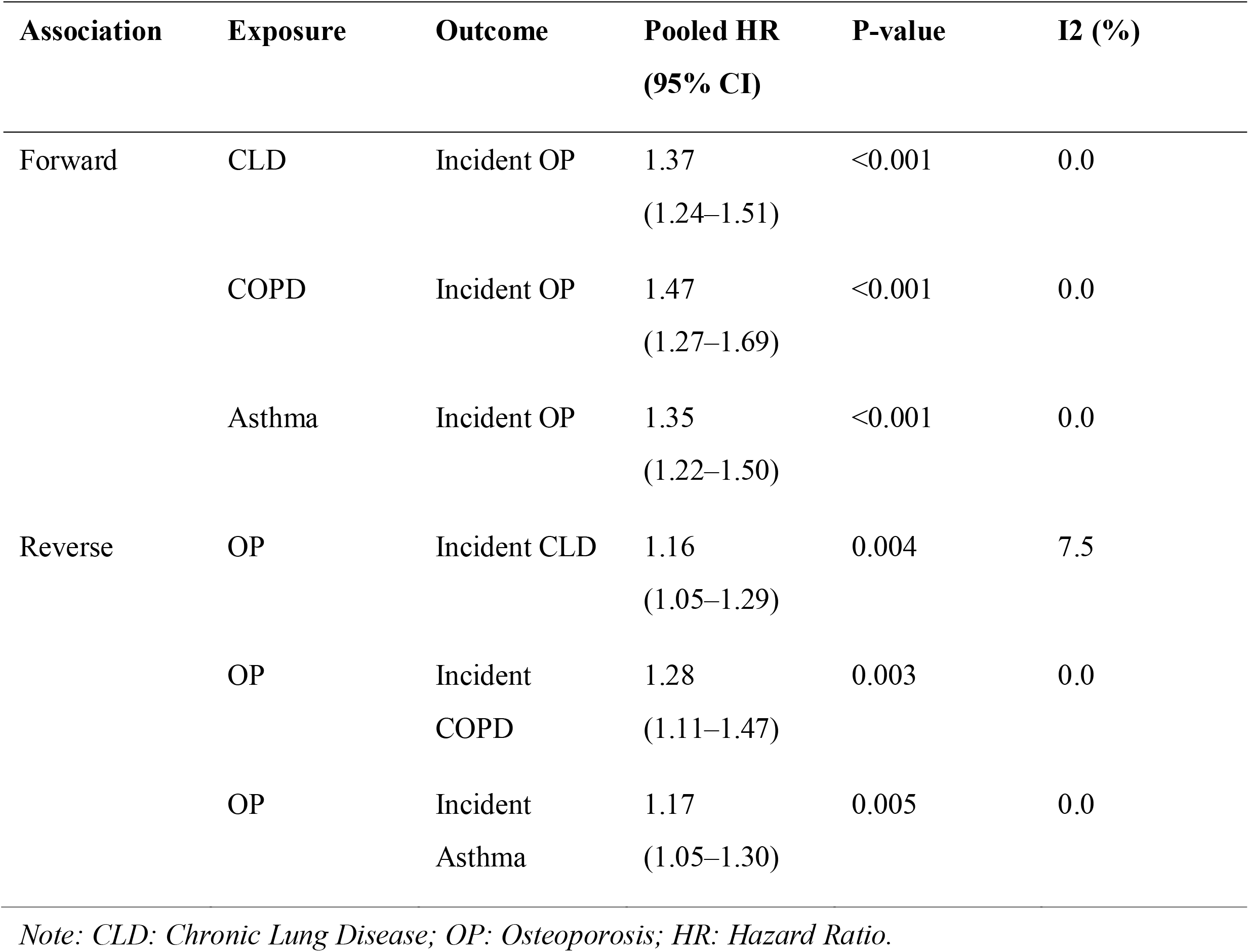
Pooled Hazard Ratios from Fixed-Effect Meta-Analysis (HRS, SHARE, ELSA)

**Figure 2.**
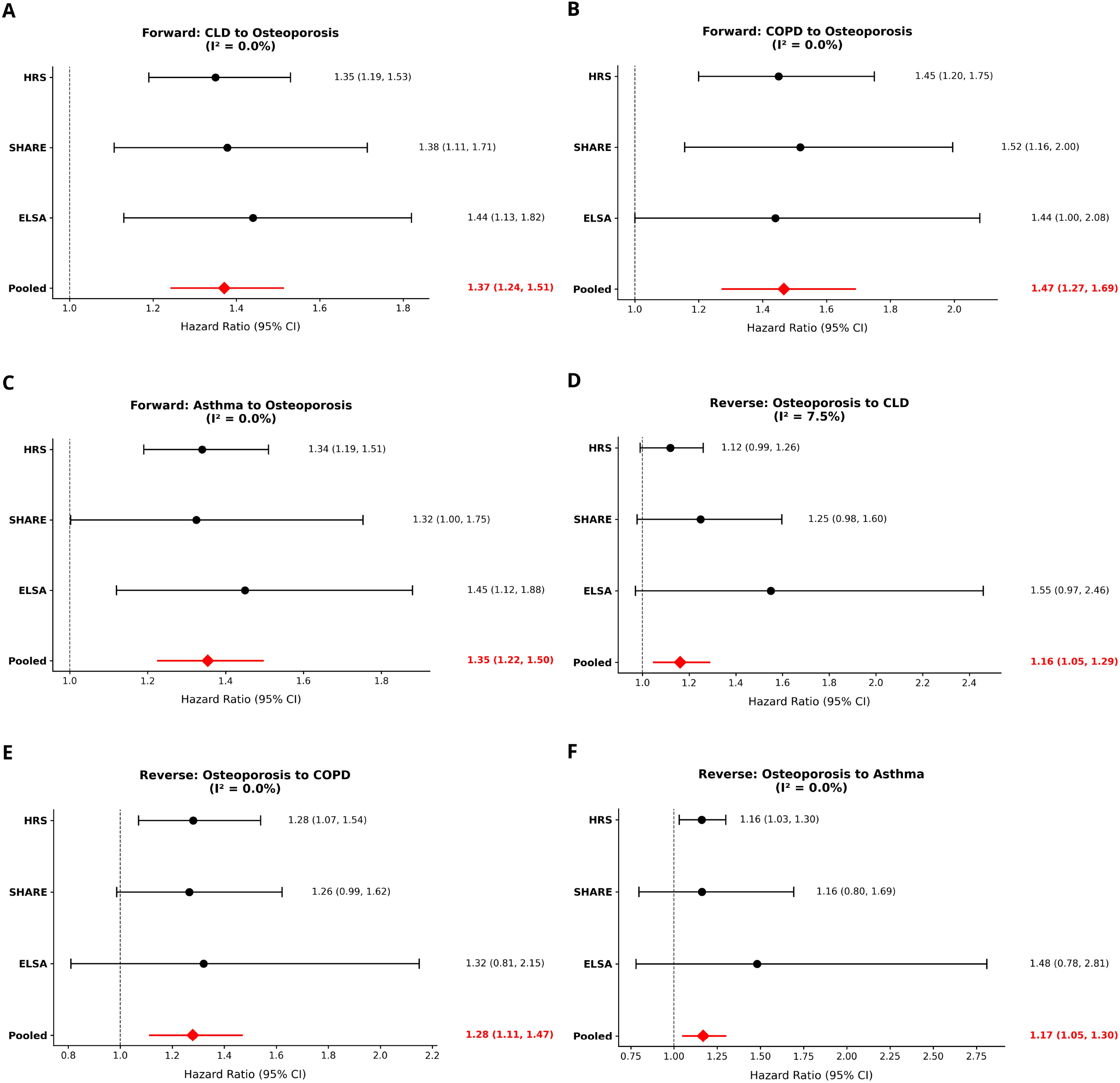
Forest Plots of the Bidirectional Association between CLD and Incident Osteoporosis. (A) CLD -> Incident OP. (B) COPD -> Incident OP. (C) Asthma -> Incident OP. (D) OP -> Incident CLD. (E) OP -> Incident COPD. (F) OP -> Incident Asthma..

For the forward association, the pooled HR for incident OP among individuals with baseline CLD was 1.37 (95% CI 1.24–1.51, P<0.001), with low heterogeneity across cohorts (I^2^=0.0%). The association was strongest for COPD (pooled HR 1.47, 95% CI 1.27–1.69, P<0.001) and slightly attenuated but still significant for asthma (pooled HR 1.35, 95% CI 1.22–1.50, P<0.001; I^2^=0.0% for both).

For the reverse association, baseline OP was associated with a 16% increased risk of incident CLD (pooled HR 1.16, 95% CI 1.05–1.29, P=0.004), with low heterogeneity (I^2^=7.5%). When examining specific CLD subtypes, OP was associated with a 28% increased risk of incident COPD (pooled HR 1.28, 95% CI 1.11–1.47, P=0.003; I^2^=0.0%) and a 17% increased risk of incident asthma (pooled HR 1.17, 95% CI 1.05–1.30, P=0.005; I^2^=0.0%).

Forest plots visually confirming these pooled estimates are presented in Figure 2 (A–F).

### Cross-sectional Comparison (LASI)

In contrast to the longitudinal findings, the cross-sectional LASI cohort yielded substantially larger effect estimates (Table 4). The OR for the association between prevalent CLD and prevalent OP was 1.78 (95% CI 1.52–2.08), representing a 30% inflation compared to the pooled longitudinal HR (1.37). The inflation was more pronounced for COPD (OR 2.04, 95% CI 1.58–2.60; 39% inflation vs. pooled HR 1.47) and for the reverse association (OP→CLD: OR 1.75, 95% CI 1.50–2.04; 51% inflation vs. pooled HR 1.16). The magnitude of effect inflation ranged from approximately 30% to 57% across all associations, empirically demonstrating the bias inherent in cross-sectional designs.

**Table 4.**
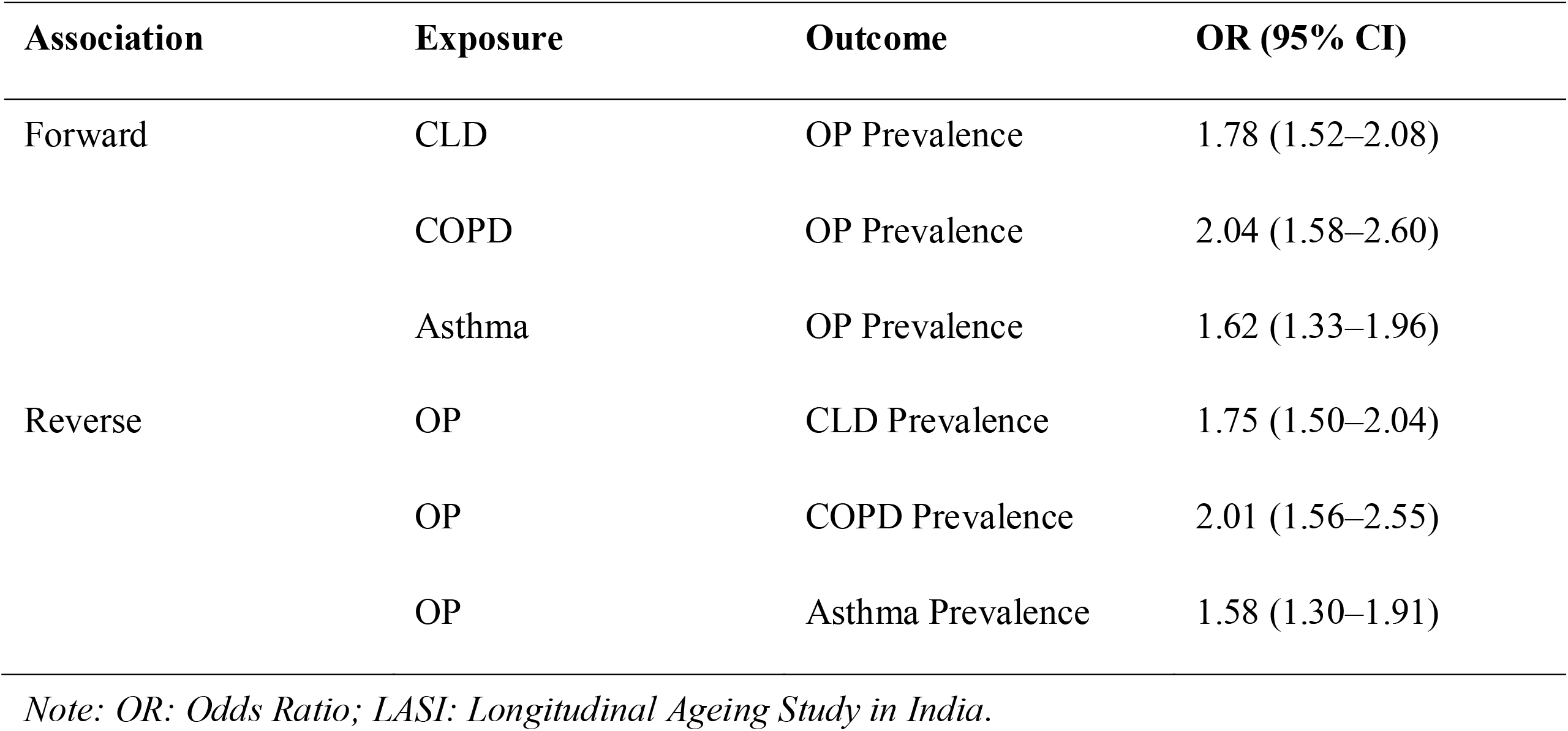
Cross-sectional Association in LASI Cohort (OR, 95% CI)

## Discussion

This multi-cohort two-stage individual participant data (IPD) meta-analysis, leveraging three large-scale international longitudinal studies (HRS, SHARE, and ELSA), provides the most robust evidence to date on the complex relationship between chronic lung disease (CLD) and osteoporosis (OP). Our core finding is the confirmation of a significant and bidirectional association between CLD (including COPD and asthma) and incident OP in aging populations. Specifically, we found that CLD significantly increases the risk of new-onset OP (pooled HR 1.37, 95% CI 1.24–1.51), and conversely, OP is an independent risk factor for the development of CLD (pooled HR 1.16, 95% CI 1.05–1.29).

This conclusion is strengthened by the consistently low heterogeneity observed across the three geographically and ethnically diverse longitudinal cohorts (I^2^ ≤7.5% for all analyses).

Our findings align with previous literature supporting the forward association (CLD → OP), which is primarily driven by systemic inflammation, shared risk factors (e.g., smoking, physical inactivity, vitamin D deficiency), and disease-specific mechanisms such as corticosteroid use in asthma and COPD [1,2,7]. However, the most critical contribution of our study lies in the confirmation of the reverse association (OP → CLD), which has yielded inconsistent results in prior single-cohort studies [8]. The use of a two-stage IPD approach allowed us to apply uniform inclusion criteria, consistent covariate definitions, and identical analytical models across cohorts, thereby minimizing methodological heterogeneity and providing more precise estimates than traditional aggregate-data meta-analyses. By pooling individual-level data, we achieved sufficient statistical power to detect this more subtle, yet clinically important, risk.

Crucially, our comparison with the cross-sectional LASI cohort revealed a profound methodological insight: the odds ratios from LASI were inflated by 30% to 57% compared to the pooled longitudinal hazard ratios. This empirical quantification underscores the inherent limitations of cross-sectional designs in chronic disease comorbidity research, where reverse causation (e.g., undiagnosed CLD leading to OP, or OP-related immobility contributing to pulmonary decline) and survival bias can severely overestimate the true effect size [9]. The substantially lower BMI and OP prevalence observed in the LASI cohort further suggest that population-specific factors, such as nutritional status and body composition, may interact with study design to influence effect estimates. These findings highlight the necessity of longitudinal designs with clear temporal sequencing when investigating causal relationships in chronic disease epidemiology.

The confirmed bidirectional link necessitates a paradigm shift toward the integrated management of CLD and OP in aging populations. From a clinical perspective, our findings suggest that patients newly diagnosed with CLD—particularly COPD—should be routinely screened for OP, and conversely, patients presenting with OP should be monitored for pulmonary symptoms and considered for lung function assessment if clinically indicated. Such integrated approaches align with emerging recommendations for multimorbidity management in older adults [10]. The mechanisms underlying the OP → CLD association, while not directly tested in this study due to data limitations, are likely mediated by several pathways. Osteoporosis-related vertebral compression fractures (VCFs) can lead to thoracic kyphosis, reduced chest wall compliance, and restrictive lung physiology, potentially contributing to the development or exacerbation of CLD symptoms [3,10]. Additionally, shared risk factors including smoking, physical inactivity, and systemic inflammation may contribute to both conditions through overlapping biological pathways [1]. Future research should prioritize studies incorporating objective measures (e.g., spirometry for CLD, dual-energy X-ray absorptiometry for OP, and spinal imaging for VCFs) to confirm these findings and elucidate underlying mechanisms. Mendelian randomization approaches using genetic instruments could further strengthen causal inference. Moreover, intervention studies are needed to determine whether integrated screening and treatment strategies improve clinical outcomes in patients with comorbid CLD and OP.

The primary strength of this study is its two-stage IPD meta-analysis design, which provides the highest level of evidence among observational study designs. By combining individual-level data from three harmonized international cohorts, we minimized the risk of single-cohort bias and maximized the generalizability of our findings to Western aging populations. The use of Cox proportional hazards models allowed for robust estimation of time-to-event data, providing strong evidence for temporality—a key requirement for causal inference. Furthermore, comprehensive adjustment for a wide range of covariates, including demographics, lifestyle factors, multiple comorbidities, and functional status, significantly reduced the potential for confounding. The consistently low heterogeneity across cohorts (I^2^ ≤7.5%) supports the robustness and consistency of our pooled estimates. Finally, the direct comparison with a large cross-sectional cohort (LASI) serves as a powerful methodological demonstration of the pitfalls of cross-sectional designs in this field and provides empirical evidence for the degree of effect size inflation that can occur when temporality is not established.

Despite these strengths, several limitations must be acknowledged. First, the diagnosis of CLD and OP across all cohorts relied on self-reported physician diagnosis, which may be subject to recall bias and misclassification compared to objective clinical measures (e.g., spirometry for CLD or dual-energy X-ray absorptiometry for OP). This limitation is particularly relevant for osteoporosis, which is often underdiagnosed in older adults due to lack of routine screening. Second, due to data unavailability, we were unable to adjust for the use of corticosteroids—a major confounder in the asthma→OP pathway—or directly test the mediating role of vertebral compression fractures in the OP→CLD pathway. Third, the exclusion of participants with missing data (complete case analysis) may introduce selection bias, potentially limiting the generalizability of our findings to the broader population. Fourth, as with all observational studies using aging cohorts, our estimates may be influenced by survivor bias—individuals with severe CLD or OP may have been lost to follow-up or deceased prior to outcome ascertainment, potentially biasing results toward the null. Fifth, the restriction to participants aged ≥50 years means our findings may not be generalizable to younger populations, in whom the etiology and progression of CLD and OP may differ. Finally, while the two-stage IPD meta-analysis strengthens causal inference relative to cross-sectional designs, as an observational study it cannot definitively prove causality, and residual confounding from unmeasured factors (e.g., genetic predisposition, environmental exposures, medication adherence) remains a possibility.

## Conclusion

This two-stage individual participant data meta-analysis of three international longitudinal cohorts provides robust evidence for a bidirectional association between chronic lung disease and incident osteoporosis in aging populations. Individuals with baseline CLD had a 37% increased risk of developing OP, while those with baseline OP had a 16% increased risk of developing CLD. The consistently low heterogeneity across cohorts supports the generalizability of these findings. Our empirical comparison with a cross-sectional cohort demonstrates that cross-sectional designs may substantially overestimate this association, underscoring the importance of longitudinal designs with clear temporal sequencing. These findings support integrated screening and management strategies for CLD and OP in older adults.

## References

1. Wang Z, Sun Y (2024) Unraveling the causality between chronic obstructive pulmonary disease and its common comorbidities using bidirectional Mendelian randomization. Eur J Med Res 29:143

2. Jones A, Fay JK, Burr M, Stone M, Hood K, Roberts G (2002) Inhaled corticosteroid effects on bone metabolism in asthma and mild chronic obstructive pulmonary disease. Cochrane Database Syst Rev 2002:CD003537

3. Bolton CE, Ionescu AA, Shiels KM, Pettit RJ, Edwards PH, Stone MD, Nixon LS, Evans WD, Griffiths TL, Shale DJ (2004) Associated loss of fat-free mass and bone mineral density in chronic obstructive pulmonary disease. Am J Respir Crit Care Med 170:1286–1293

4. Penedones A, Mendes D, Alves C, Filipe A, Oliveira T, Batel-Marques F (2024) Relationship Between Chronic Obstructive Pulmonary Disease and Osteoporosis: A Scoping Review. COPD 21:2356510

5. Rodriguez-Gomez I, Gray SR, Ho FK, et al. (2022) Osteoporosis and Its Association With Cardiovascular Disease, Respiratory Disease, and Cancer: Findings From the UK Biobank Prospective Cohort Study. Mayo Clin Proc 97:110–121

6. Mohammednoor M, Mubarak Osman AME, Hamad S, Abdelhalim Ismail FI, Elsharief Mohammed Elamien MEY, Mohamed Gamareldin EE, Rahman MH (2025) Impact of Inhaled Corticosteroids on Osteoporosis in Chronic Obstructive Pulmonary Disease (COPD): A Systematic Review. Cureus 17:e89201

7. Huang HF, Hsu CW, Hsieh FT, Liao KM (2025) Evaluating inhaled corticosteroids’ impact on osteoporosis and fracture risk in COPD patients: a real-world evidence-based systematic review and meta-analysis. Front Med (Lausanne) 12:1503475

8. Tu Y, Miao J, Wu Q, Lu K, Ren R, Lin C, Wang X, Jin H (2025) Obstructive sleep apnea and osteoporosis: A bidirectional two-sample mendelian randomization analysis. Respir Med 242:108090

9. Negewo NA, Gibson PG, McDonald VM (2015) COPD and its comorbidities: Impact, measurement and mechanisms. Respirology 20:1160–1171

10. Mirza S, Clay RD, Koslow MA, Scanlon PD (2018) COPD Guidelines: A Review of the 2018 GOLD Report. Mayo Clin Proc 93:1488–1502

