## supplemental table 1-7 for "The Robust Bidirectional Association Between Chronic Lung Disease and Incident Osteoporosis: A Two-Stage Individual Participant Data Meta-Analysis of Three International Longitudinal Cohorts (HRS, SHARE, and ELSA)"

**Supplementary table 1. Baseline Characteristics by CLD Subgroups (HRS)**

| Variable | Total (N=17,996) | No CLD (N=14,463) | CLD (N=3,533) | COPD (N=612) | Asthma (N=1,838) | P-value |
| --- | --- | --- | --- | --- | --- | --- |
| Age (years), Mean (SD) | 66.99 (10.61) | 66.82 (10.68) | 67.67 (10.33) | 69.39 (10.10) | 66.30 (10.27) | <0.001 |
| BMI (kg/m²), Mean (SD) | 28.61 (6.19) | 28.32 (5.84) | 29.76 (7.35) | 28.60 (7.08) | 30.17 (6.92) | <0.001 |
| Gender (Female), n (%) | 10453 (58.1%) | 8106 (56.0%) | 2347 (66.4%) | 364 (59.5%) | 1307 (71.1%) | <0.001 |
| Marital Status (Married), n (%) | 11250 (62.5%) | 9282 (64.2%) | 1968 (55.7%) | 335 (54.7%) | 1072 (58.3%) | <0.001 |
| Education Level, n (%) |  |  |  |  |  | <0.001 |
| - Less than high school | 4115 (22.9%) | 3077 (21.3%) | 1038 (29.4%) | 173 (28.3%) | 483 (26.3%) |  |
| - High school | 5154 (28.6%) | 4157 (28.7%) | 997 (28.2%) | 196 (32.0%) | 475 (25.8%) |  |
| - College and above | 8727 (48.5%) | 7229 (50.0%) | 1498 (42.4%) | 243 (39.7%) | 880 (47.9%) |  |
| Smoking Status (Smoker), n (%) | 10177 (56.6%) | 7845 (54.2%) | 2332 (66.0%) | 473 (77.3%) | 962 (52.3%) | <0.001 |
| Drinking Status (Drinker), n (%) | 9898 (55.0%) | 8247 (57.0%) | 1651 (46.7%) | 283 (46.2%) | 922 (50.2%) | <0.001 |
| Physical Activity, n (%) |  |  |  |  |  | <0.001 |
| - Low | 8344 (46.4%) | 6325 (43.7%) | 2019 (57.1%) | 347 (56.7%) | 931 (50.7%) |  |
| - Moderate | 5146 (28.6%) | 4215 (29.1%) | 931 (26.4%) | 170 (27.8%) | 514 (28.0%) |  |
| - Vigorous | 4506 (25.0%) | 3923 (27.1%) | 583 (16.5%) | 95 (15.5%) | 393 (21.4%) |  |
| Comorbidities, n (%) |  |  |  |  |  |  |
| - Hypertension | 10706 (59.5%) | 8183 (56.6%) | 2523 (71.4%) | 399 (65.2%) | 1309 (71.2%) | <0.001 |
| - Diabetes | 4159 (23.1%) | 3097 (21.4%) | 1062 (30.1%) | 167 (27.3%) | 554 (30.1%) | <0.001 |
| - Heart Disease | 4166 (23.1%) | 2939 (20.3%) | 1227 (34.7%) | 228 (37.3%) | 497 (27.0%) | <0.001 |
| - Stroke | 1462 (8.1%) | 1029 (7.1%) | 433 (12.3%) | 83 (13.6%) | 179 (9.7%) | <0.001 |
| - Cancer | 2579 (14.3%) | 1962 (13.6%) | 617 (17.5%) | 129 (21.1%) | 264 (14.4%) | <0.001 |
| - Arthritis | 10182 (56.6%) | 7547 (52.2%) | 2635 (74.6%) | 461 (75.3%) | 1303 (70.9%) | <0.001 |
| Depression Risk, n (%) | 2791 (15.5%) | 1840 (12.7%) | 951 (26.9%) | 146 (23.9%) | 445 (24.2%) | <0.001 |
| ADL score, Mean (SD) | 0.30 (0.84) | 0.22 (0.72) | 0.63 (1.16) | 0.47 (1.00) | 0.58 (1.15) | <0.001 |
| Osteoporosis, n (%) | 2423 (13.5%) | 1676 (11.6%) | 747 (21.1%) | 116 (19.0%) | 338 (18.4%) | <0.001 |

### Supplementary table 2. Baseline Characteristics by CLD Subgroups (SHARE)

| Variable | Total | No CLD | CLD | COPD | Asthma | P-value |
| --- | --- | --- | --- | --- | --- | --- |
|  | (N=29,151) | (N=26,683) | (N=2,468) | (N=1,455) | (N=1,323) |  |
| Age (years), Mean (SD) | 64.58 (10.09) | 64.37 (10.04) | 66.92 (10.31) | 68.11 (10.22) | 65.84 (10.27) | <0.001 |
| BMI (kg/m²), Mean (SD) | 26.41 (4.30) | 26.36 (4.24) | 26.94 (4.92) | 26.84 (4.94) | 27.03 (4.96) | <0.001 |
| Gender (Female), n (%) | 15,814 (54.2%) | 14,529 (54.5%) | 1,285 (52.1%) | 707 (48.6%) | 757 (57.2%) | <0.001 |
| Marital Status (Married), n (%) | 23,022 (79.0%) | 21,208 (79.5%) | 1,814 (73.5%) | 1,054 (72.4%) | 986 (74.5%) | <0.001 |
| Education Level, n (%) |  |  |  |  |  |  |
| - Less than high school | 15,200 (52.1%) | 13,696 (51.3%) | 1,504 (60.9%) | 938 (64.5%) | 765 (57.8%) | <0.001 |
| - High school | 8,560 (29.4%) | 7,952 (29.8%) | 608 (24.6%) | 337 (23.2%) | 344 (26.0%) | <0.001 |
| - College and above | 5,391 (18.5%) | 5,035 (18.9%) | 356 (14.4%) | 180 (12.4%) | 214 (16.2%) | <0.001 |
| Smoking Status (Smoker), n (%) | 13,654 (46.8%) | 12,269 (46.0%) | 1,385 (56.1%) | 886 (60.9%) | 686 (51.9%) | <0.001 |
| Drinking Status (Drinker), n (%) | 3,067 (10.5%) | 2,798 (10.5%) | 269 (10.9%) | 149 (10.2%) | 153 (11.6%) | 0.549 |
| Physical Activity, n (%) |  |  |  |  |  |  |
| - Low | 29,151 (100.0%) | 26,683 (100.0%) | 2,468 (100.0%) | 1,455 (100.0%) | 1,323 (100.0%) | 1.000 |
| - Moderate | 0 (0.0%) | 0 (0.0%) | 0 (0.0%) | 0 (0.0%) | 0 (0.0%) | 1.000 |
| - Vigorous | 0 (0.0%) | 0 (0.0%) | 0 (0.0%) | 0 (0.0%) | 0 (0.0%) | 1.000 |
| Comorbidities, n (%) |  |  |  |  |  |  |
| - Hypertension | 9,416 (32.3%) | 8,432 (31.6%) | 984 (39.9%) | 588 (40.4%) | 532 (40.2%) | <0.001 |
| - Diabetes | 3,010 (10.3%) | 2,673 (10.0%) | 337 (13.7%) | 213 (14.6%) | 168 (12.7%) | <0.001 |
| - Heart Disease | 3,621 (12.4%) | 3,088 (11.6%) | 533 (21.6%) | 353 (24.3%) | 263 (19.9%) | <0.001 |
| - Stroke | 1,090 (3.7%) | 962 (3.6%) | 128 (5.2%) | 83 (5.7%) | 65 (4.9%) | <0.001 |
| - Cancer | 1,547 (5.3%) | 1,372 (5.1%) | 175 (7.1%) | 122 (8.4%) | 76 (5.7%) | <0.001 |
| - Arthritis | 5,524 (18.9%) | 4,751 (17.8%) | 773 (31.3%) | 501 (34.4%) | 412 (31.1%) | <0.001 |
| Depression Risk, n (%) | 7,536 (25.9%) | 6,545 (24.5%) | 991 (40.2%) | 616 (42.3%) | 533 (40.3%) | <0.001 |
| ADL score, Mean (SD) | 0.20 (0.71) | 0.18 (0.68) | 0.39 (0.96) | 0.46 (1.04) | 0.36 (0.90) | <0.001 |
| Osteoporosis, n (%) | 2,266 (7.8%) | 1,958 (7.3%) | 308 (12.5%) | 200 (13.7%) | 158 (11.9%) | <0.001 |

**Supplementary table 3. Baseline Characteristics by CLD Subgroups (ELSA)**

| Variable | Total (N=5806) | No CLD (N=4878) | CLD (N=928) | COPD (N=345) | Asthma (N=688) | P-value |
| --- | --- | --- | --- | --- | --- | --- |
| Age (years), Mean (SD) | 63.42 (9.20) | 63.31 (9.22) | 64.00 (9.05) | 66.17 (9.02) | 63.29 (8.92) | <0.001 |
| BMI (kg/m2), Mean (SD) | 27.94 (4.87) | 27.89 (4.78) | 28.18 (5.34) | 27.65 (5.34) | 28.49 (5.39) | 0.005 |
| Gender (Female), n (%) | 3070 (52.9%) | 2556 (52.4%) | 514 (55.4%) | 167 (48.4%) | 408 (59.3%) | <0.001 |
| Marital Status (Married), n (%) | 4291 (73.9%) | 3635 (74.5%) | 656 (70.7%) | 221 (64.1%) | 499 (72.5%) | <0.001 |
| Education Level, n (%) |  |  |  |  |  | <0.001 |
| - Less than high school | 2572 (44.3%) | 2100 (43.1%) | 472 (50.9%) | 216 (62.6%) | 324 (47.1%) |  |
| - High school | 1162 (20.0%) | 994 (20.4%) | 168 (18.1%) | 58 (16.8%) | 128 (18.6%) |  |
| - College and above | 2072 (35.7%) | 1784 (36.6%) | 288 (31.0%) | 71 (20.6%) | 236 (34.3%) |  |
| Smoking Status (Smoker), n (%) | 3666 (63.1%) | 3025 (62.0%) | 641 (69.1%) | 272 (78.8%) | 449 (65.3%) | <0.001 |
| Drinking Status (Drinker), n (%) | 5231 (90.1%) | 4418 (90.6%) | 813 (87.6%) | 287 (83.2%) | 610 (88.7%) | <0.001 |
| Physical Activity, n (%) |  |  |  |  |  | <0.001 |
| - Low | 747 (12.9%) | 562 (11.5%) | 185 (19.9%) | 97 (28.1%) | 121 (17.6%) |  |
| - Moderate | 2643 (45.5%) | 2212 (45.3%) | 431 (46.4%) | 170 (49.3%) | 316 (45.9%) |  |
| - Vigorous | 2416 (41.6%) | 2104 (43.1%) | 312 (33.6%) | 78 (22.6%) | 251 (36.5%) |  |
| Comorbidities, n (%) |  |  |  |  |  |  |
| - Hypertension | 2308 (39.8%) | 1894 (38.8%) | 414 (44.6%) | 155 (44.9%) | 311 (45.2%) | 0.002 |
| - Diabetes | 433 (7.5%) | 372 (7.6%) | 61 (6.6%) | 27 (7.8%) | 41 (6.0%) | 0.443 |
| - Heart Disease | 946 (16.3%) | 766 (15.7%) | 180 (19.4%) | 87 (25.2%) | 123 (17.9%) | <0.001 |
| - Stroke | 212 (3.7%) | 174 (3.6%) | 38 (4.1%) | 14 (4.1%) | 28 (4.1%) | 0.923 |
| - Cancer | 412 (7.1%) | 342 (7.0%) | 70 (7.5%) | 29 (8.4%) | 48 (7.0%) | 0.505 |
| - Arthritis | 1968 (33.9%) | 1596 (32.7%) | 372 (40.1%) | 146 (42.3%) | 278 (40.4%) | <0.001 |
| Depression Risk, n (%) | 783 (13.5%) | 591 (12.1%) | 192 (20.7%) | 92 (26.7%) | 135 (19.6%) | <0.001 |
| ADL score, Mean (SD) | 0.31 (0.79) | 0.27 (0.75) | 0.51 (0.98) | 0.70 (1.09) | 0.48 (0.98) | <0.001 |
| Osteoporosis, n (%) | 285 (4.9%) | 205 (4.2%) | 80 (8.6%) | 39 (11.3%) | 58 (8.4%) | <0.001 |

**Supplementary table 4. Baseline Characteristics by CLD Subgroups (LASI)**

| Variable | Total (N=52,951) | No CLD (N=49,678) | CLD (N=3,273) | COPD (N=972) | Asthma (N=2,122) | P-value |
| --- | --- | --- | --- | --- | --- | --- |
| Age (years), Mean (SD) | 62.80 (9.39) | 62.80 (9.39) | 65.89 (9.88) | 65.07 (9.58) | 66.16 (9.97) | <0.001 |
| BMI (kg/m²), Mean (SD) | 22.67 (4.74) | 22.67 (4.74) | 21.93 (5.08) | 22.49 (5.25) | 21.72 (4.99) | <0.001 |
| Gender (Female), n (%) | 28,068 (53.0%) | 26,504 (53.4%) | 1,564 (47.8%) | 451 (46.4%) | 1,021 (48.1%) | <0.001 |
| Marital Status (Married), n (%) | 37,658 (71.1%) | 35,441 (71.3%) | 2,217 (67.7%) | 672 (69.1%) | 1,434 (67.6%) | <0.001 |
| Education Level, n (%) |  |  |  |  |  | <0.001 |
| - Less than high school | 39,352 (74.3%) | 36,774 (74.0%) | 2,578 (78.8%) | 696 (71.6%) | 1,723 (81.2%) |  |
| - High school | 11,145 (21.0%) | 10,527 (21.2%) | 618 (18.9%) | 237 (24.4%) | 354 (16.7%) |  |
| - College and above | 2,454 (4.6%) | 2,377 (4.8%) | 77 (2.4%) | 39 (4.0%) | 45 (2.1%) |  |
| Smoking Status (Smoker), n (%) | 10,035 (19.0%) | 9,159 (18.5%) | 876 (26.8%) | 271 (28.0%) | 556 (26.4%) | <0.001 |
| Drinking Status (Drinker), n (%) | 9,430 (17.8%) | 8,795 (17.8%) | 635 (19.4%) | 177 (18.3%) | 416 (19.7%) | 0.029 |
| Physical Activity, n (%) |  |  |  |  |  | <0.001 |
| - Low | 22,701 (42.9%) | 21,045 (42.4%) | 1,656 (50.6%) | 497 (51.1%) | 1,062 (50.0%) |  |
| - Moderate | 18,630 (35.2%) | 17,558 (35.3%) | 1,072 (32.8%) | 298 (30.7%) | 721 (34.0%) |  |
| - Vigorous | 11,620 (21.9%) | 11,075 (22.3%) | 545 (16.7%) | 177 (18.2%) | 339 (16.0%) |  |
| Comorbidities, n (%) |  |  |  |  |  |  |
| - Hypertension | 16,653 (31.4%) | 15,316 (30.8%) | 1,337 (40.8%) | 432 (44.4%) | 828 (39.0%) | <0.001 |
| - Diabetes | 7,529 (14.2%) | 6,973 (14.0%) | 556 (17.0%) | 184 (18.9%) | 349 (16.4%) | <0.001 |
| - Heart Disease | 2,174 (4.1%) | 1,897 (3.8%) | 277 (8.5%) | 102 (10.5%) | 152 (7.2%) | <0.001 |
| - Stroke | 1,109 (2.1%) | 1,016 (2.0%) | 93 (2.8%) | 30 (3.1%) | 57 (2.7%) | 0.017 |
| - Cancer | 380 (0.7%) | 348 (0.7%) | 32 (1.0%) | 14 (1.4%) | 14 (0.7%) | 0.004 |
| - Arthritis | 6,698 (12.6%) | 6,078 (12.2%) | 620 (18.9%) | 190 (19.5%) | 384 (18.1%) | <0.001 |
| Depression Risk, n (%) | 22,648 (42.8%) | 21,020 (43.4%) | 1,628 (51.3%) | 493 (52.6%) | 1,037 (50.9%) | <0.001 |
| ADL score, Mean (SD) | 0.29 (0.93) | 0.28 (0.91) | 0.48 (1.18) | 0.41 (1.13) | 0.48 (1.18) | <0.001 |
| Osteoporosis, n (%) | 2,091 (3.9%) | 1,842 (3.7%) | 249 (7.6%) | 86 (8.8%) | 148 (7.0%) | <0.001 |

**Supplementary table 5. Bidirectional Association between CLD and Osteoporosis (HRS)**

| Analysis Type | Exposure | Outcome | HR (95% CI) | P-value |
| --- | --- | --- | --- | --- |
| Forward Analysis | CLD | Incident Osteoporosis | 1.35 (1.19 - 1.53) | <0.001 |
|  | COPD | Incident Osteoporosis | 1.45 (1.20 - 1.75) | <0.001 |
|  | Asthma | Incident Osteoporosis | 1.34 (1.19 - 1.51) | <0.001 |
| Reverse Analysis | Osteoporosis | Incident CLD | 1.12 (0.99 - 1.26) | 0.058 |
|  | Osteoporosis | Incident COPD | 1.28 (1.07 - 1.54) | 0.008 |
|  | Osteoporosis | Incident Asthma | 1.16 (1.03 - 1.30) | 0.014 |

Note: Models were adjusted for age, gender, marital status, education, income, smoking, drinking, BMI, physical activity, hypertension, diabetes, heart disease, stroke, cancer, arthritis, depression risk, and ADL score.

**Supplementary table 6. Bidirectional Association between CLD and Osteoporosis (SHARE)**

| Analysis Type | Exposure | Outcome | HR (95% CI) | P-value |
| --- | --- | --- | --- | --- |
| Forward Analysis | CLD | Osteoporosis | 1.378 (1.107 - 1.714) | 0.004 |
| Forward Analysis | COPD | Osteoporosis | 1.518 (1.156 - 1.995) | 0.003 |
| Forward Analysis | Asthma | Osteoporosis | 1.325 (1.002 - 1.753) | 0.049 |
| Reverse Analysis | Osteoporosis | CLD | 1.249 (0.977 - 1.597) | 0.076 |
| Reverse Analysis | Osteoporosis | COPD | 1.265 (0.986 - 1.622) | 0.065 |
| Reverse Analysis | Osteoporosis | Asthma | 1.161 (0.797 - 1.692) | 0.436 |

Note: Models were adjusted for age, gender, marital status, education, income, smoking, drinking, BMI, physical activity, hypertension, diabetes, heart disease, stroke, cancer, arthritis, depression risk, and ADL score.

**Supplementary table 7. Bidirectional Association between CLD and Osteoporosis (ELSA)**

| Analysis Type | Exposure | Outcome | HR (95% CI) | P-value |
| --- | --- | --- | --- | --- |
| Forward Analysis | CLD | Incident Osteoporosis | 1.44 (1.13 - 1.82) | 0.003 |
|  | COPD | Incident Osteoporosis | 1.44 (1.00 - 2.08) | 0.051 |
|  | Asthma | Incident Osteoporosis | 1.45 (1.12 - 1.88) | 0.005 |
| Reverse Analysis | Osteoporosis | Incident CLD | 1.55 (0.97 - 2.46) | 0.064 |
|  | Osteoporosis | Incident COPD | 1.32 (0.81 - 2.15) | 0.264 |
|  | Osteoporosis | Incident Asthma | 1.48 (0.78 - 2.81) | 0.230 |

Note: Models were adjusted for age, gender, marital status, education, income, smoking, drinking, BMI, physical activity, hypertension, diabetes, heart disease, stroke, cancer, arthritis, depression risk, and ADL score.
